# MetDecode: methylation-based deconvolution of cell-free DNA for non-invasive multi-cancer typing

**DOI:** 10.1101/2023.12.29.23300371

**Authors:** Dhanya Sudhakaran, Stefania Tuveri, Antoine Passemiers, Tatjana Jatsenko, Tina Laga, Kevin Punie, Sabine Tejpar, An Coosemans, Els Van Nieuwenhuysen, Dirk Timmerman, Giuseppe Floris, Anne-Sophie Van Rompuy, Xavier Sagaert, Antonia Testa, Daniela Ficherova, Daniele Raimondi, Frederic Amant, Liesbeth Lenaerts, Yves Moreau, Joris R. Vermeesch

## Abstract

Cell-free DNA (cfDNA) mediated early cancer detection is based on detecting alterations in the cfDNA components. However, the underlying pathology can usually not be readily identified. We built a reference atlas based on the methylome of multiple cancer and blood-cell types and developed MetDecode, an epigenetic signature-based deconvolution algorithm. MetDecode accurately estimates the tumour proportion in *in-silico* mixtures and identifies the tissue of origin in 81.25% cfDNA samples from cancer patients. This method will complement cancer screening programs and guide clinical follow-up.

## Background

Cell-free DNA (cfDNA) is present as DNA fragments floating in the blood. The fragments, mainly derived from dying cells, contain the genomic and epigenetic signatures of the cells of origin [1]. When cancer is present, a fraction of the cfDNA can be derived from tumour cells, defined as circulating-tumour DNA (ctDNA) [2]. ctDNA is now being widely explored as a non-invasive biomarker for cancer screening and diagnosis [3]. A major focus is on the detection of cancer-specific single nucleotide mutations and copy number alterations (CNAs). Whereas somatic mutations are usually identified by targeted sequencing, the detection of CNAs is done by genome-wide sequencing of the cfDNA [4,5]. Though somatic mutations can be tumour-specific, their application for cancer detection and screening is hampered by the low variant allele frequency at the early stages of the disease [5,6]. Alternatively, genome-wide detection of tumour-associated CNAs in cfDNA has been shown to allow cancer detection, also in population screening settings with low-pass sequencing [7–10].

CfDNA-based screening for cancers can often detect the presence of abnormal signals indicative of a cancer, but not its origin or cancer type. Especially for metastatic disease of unknown primary, profiles do not readily allow the identification of the tissue of origin (TOO) albeit this would be of clinical value [11]. When performing non-invasive prenatal screening for foetal chromosomal aneuploidies, incidental occult maternal malignancies can be detected without insights into the origin [10,12,13]. If the TOO or cancer type could be deduced from cfDNA analysis, this would tremendously speed up the diagnosis and start of treatment, hence streamlining subsequent clinical follow-up, reducing costs and minimising the need for extensive radiologic imaging [14]. For patients, this might reduce the anxiety associated with a positive screening test outcome.

Methods to deduce the origin of cfDNA fragments have been based on epigenetic markers such as nucleosome positioning, fragmentation and methylation profiles [15–17]. These profiles are tissue and cell-type-specific [18], offering the possibility to identify the different components of the cfDNA pool, alongside an estimation of the relative proportion of each of them. Tumour-associated methylation changes have been described during cancer initiation and progression. Hence, they are promising markers for early cancer TOO identification [19].

Recently, several algorithms have been developed to deconvolute the plasma cfDNA composition based on methylation profiles. Typically, reference atlases consisting of either normal tissues or cell-type-specific methylation markers are used to identify tissue-specific methylation signals [14,19–26]. Although each method has its specific merits, it also has its limitations. For instance, none of the methods deconvolutes multiple cancer tissues. Also, most methods do not consider missing variables due to the incompleteness of the atlas [22,28] or operate in a reference-free fashion [26]. However, cfDNA mixtures are more complex and could carry fragments from tissues not represented in the atlas.

To address these limitations, we developed an alternative reference-based deconvolution method, named MetDecode. The method builds on gradient-based optimization and extends existing methods by simultaneously modelling the presence of noise and the lack of comprehensiveness of the reference atlas in a deterministic and lowly-parametrized fashion. We used in-house sequenced or publicly available tumour ssamples to build a reference atlas of tissue-specific methylation markers for four different cancer tissues, namely breast, ovarian, cervical and colorectal cancer and combined it with white blood cell (WBC)-derived entities. The reference atlas is subsequently exploited by MetDecode to estimate the contribution of each atlas entity. Additionally, the reference atlas is extended with unknown methylation patterns learnt on-the-fly from cfDNA methylation profiles to account for missing data. This method could complement cancer screening programs to direct clinical follow-up to the right cancer type and will expedite treatment.

## Methods

### Plasma cfDNA and genomic DNA collection and extraction

Peripheral blood was collected in Roche cell-free DNA blood collection tubes® (Roche, Switzerland) or a Streck Cell-Free BCT® (Streck, USA) and extracted as described previously [7]. Archived [7] and prospectively collected plasma cfDNA samples of healthy individuals were included as control samples (18-90 years old). We included only individuals without cancer and no known autoimmune condition to exclude the introduction of confounding factors to the analysis, as both pathological conditions can influence the shedding and the composition of cfDNA [2,29]. Archived plasma cfDNA was also obtained from patients with a known diagnosis of breast, colorectal or ovarian cancer (mean age: 61.88 years old). Treatment-naïve formalin-fixed paraffin-embedded (FFPE) tumour biopsies were collected. Genomic DNA (gDNA) was extracted from the FFPE tumour biopsies as well as from WBC from healthy subjects or patients with a diagnosis of breast, colorectal, cervical or ovarian cancer using the QIAamp DNA FFPE Tissue Kit or the DNeasy Blood & Tissue Kits (Qiagen, Hilden, Germany), respectively. The extracted gDNA was fragmented using Covaris M220 before library preparation (Covaris Inc., Woburn, MA, USA). The study was approved by the ethical committee of the University Hospitals Leuven (study protocols S62285, S62795, S63983, S66450, S59207 and S51375).

### Complete blood count

Advia 2120 hemacytometer was used to perform the complete blood count (CBC) analysis on whole blood following manufacturer’s instructions.

### Whole-genome methylation sequencing and data analysis

cfDNA and gDNA extracted from FFPE tumour biopsies or WBC was subjected to whole-genome DNA methylation sequencing using the NEBNext Enzymatic Methyl-seq kit (New England Biolabs, Ipswich, MA, USA) following manufacturer’s instructions. Enzymatic conversion was preferred over bisulfite conversion for methylation analysis, avoiding fragmentation and loss of DNA in the process [28,30]. In addition, for gDNA from cervical and ovarian FFPE tumour biopsies that were used to build the reference atlas, bisulfite conversion was performed, to be consistent with the method used for the remaining samples in the atlas. Libraries were prepared with the same kit, thereby replacing the enzymatic conversion reactions with the bisulfite treatment using EZ-96 DNA Methylation-Direct MagPrep (Zymo Research, Irvine, CA, USA). The conversion efficiency was evaluated by spiking unmethylated Lambda DNA in one sample per batch, irrespective of the conversion method used. Libraries were quantified using Qubit dsDNA high-sensitivity assay kit and Qubit 3.0 fluorometer (Thermo Fisher Scientific, Waltham, MA, USA). Libraries were sequenced on NovaSeq 6000 S4 flowcell (Illumina, San Diego, CA, USA) generating PE150bp reads at an average depth of 15X. The data after demultiplexing was quality checked and trimmed using fastp (v0.20) and then aligned to human genome hg38 using bwa-meth (v0.2.2). Deduplication was done using Picard (v2.20.3) and methylation calling via MethylDackel (v0.5.1). The tumour fraction in the cfDNA samples was calculated using ichorCNA [31]

### Generation of a DNA methylation marker atlas for multiple blood cell types and tumour tissues

A DNA methylation marker atlas, covering markers for 6 tumour tissues and 7 blood cell types was generated solely using whole-genome bisulfite sequencing (WGBS) data. From public repositories, we downloaded genome-wide CpG site methylation ratios for B cells, CD4+ T cells, CD8+ T cells, natural killer cells, monocytes, neutrophils and erythroblasts (BLUEPRINT [32], GSE186458), and for breast invasive carcinoma, colon adenocarcinoma and rectal adenocarcinoma tissues (The Cancer Genome Atlas [33] (Supplementary Table 1, Additional File 2). WGBS data for high-grade serous ovarian carcinoma, cervical adenocarcinoma and cervical squamocellular carcinoma were generated in-house from FFPE samples. Available samples (n=2-7) per tissue/cell type were merged after removing highly variant CpG site which resulted in a combined atlas entity for every tissue as explained in the Additional File 1. Using these combined atlas entities, the sites which were uniquely methylated in one tissue type, with at least 30% difference between the absolute methylation value in that tissue versus the rest, were extracted using R scripts and extended to cover a region with a minimum of 4 CpGs (if the sites were within 500 bp). Once the start and end coordinates of the marker regions were obtained, the total number of reads and the number of methylated reads in these regions for every tissue/cell-type in the atlas, namely *D*^(*atlas*)^ and *M*^(*atlas*)^, were obtained using custom scripts. The same was also extracted for the samples to be deconvoluted (*D*^(*cfdna*)^, *M*^(*cfdna*)^) and then used as input for the deconvolution algorithm.

### Deconvolution algorithm

We created a methylation-based reference atlas, composed of two matrices *D*^(*atlas*)^ and *M*^(*atlas*)^, where 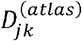 is the total CpG count for atlas entity *j* and marker region *k* ^(*atlas*)^, and 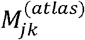 the corresponding methylated CpG count. Because each CpG site can be spanned by multiple reads, it may contribute multiple times to the same count. Therefore, these values must not be confused with read counts. We also provided the algorithm with two other input matrices, *D*^(*cfdna*)^ and *M*^(*cfdna*)^, representing the cfDNA mixtures. Our algorithm has been designed to infer a matrix of cell type contributions, where j is the estimated proportion of cell type *j* to cfDNA profile. *A* was found by minimizing a weighted mean squared error between the methylation ratios of cfDNA samples and the ratios of convoluted atlas entities.

Marker region *k* in sample *i* was re-weighted by 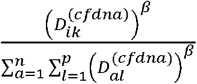 to better reflect the confidence in the estimation of the methylation 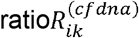. *β* is a hyper-parameter (default=1) controlling the importance given by the end user to the coverage. To account for the presence of unknown cell types missing from the reference atlas, we extended the atlas with estimates of missing cell types. When the number of cfDNA samples is largely greater than the atlas size, the methylation patterns of these unknown contributors can be learned from the data directly. The assumed number of unknown cell types was defined as a hyper-parameter (default = 1). We accounted for the unknown contributors in the cfDNA mixture by appending extra rows to the *D*^(*cfdna*)^ and *M*^(*cfdna*)^ matrices. Methylation ratio matrices *R*^(*cfdna*)^ and *R*^(*atlas*)^ were computed from the corresponding read count matrices.

*R*^(*cfdna*)^ was next deconvoluted using non-negative least squares (NNLS) algorithm and reference matrix *R*^(*atlas*)^, the residuals were used to define the missing contributor and extend *D*^(*atlas*)^, *R*^(*atlas*)^ and *M*^(*atlas*)^ by one row each. This procedure was repeated *h* times. A more technical description of the algorithm is provided in Additional File 1.

### Evaluation metric

Pearson Correlation Coefficient and mean squared error (MSE) were used to evaluate the reliability of MetDecode estimations. We evaluated the accuracy of multiclass cancer TOO prediction as 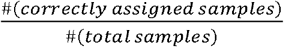. P-values were considered significant when <0.001.

## Results

### Creation of a reference atlas and tissue-specific epigenetic marker selection

To enable the deconvolution of a methylome into its potential contributors by assigning the relative proportion to a specific tissue type, a methylation reference atlas with 13 entities was created. We included methylome data from seven cells of hematopoietic origin which are the most represented in plasma cfDNA [22,34] as well as methylome data from six different tumours. The tumour tissues included breast cancer, ovarian cancer, colon adenocarcinoma, rectum adenocarcinoma, cervical adenocarcinoma and cervical squamous cell carcinoma. These cancers were selected to serve as a proof-of-concept. The seven cell types from the haematopoietic lineage included neutrophils, monocytes, erythroblasts, natural killer, B cells, CD4+ T cells and CD8+ T cells. Tumour methylome data was downloaded from TCGA for five breast invasive carcinomas from different subtypes (n=1 luminal A, n=1 luminal B, n=1 basal-like; n=2 HER2), two rectum adenocarcinoma and two colon adenocarcinoma (Supplementary Table 1, Additional File 2). Since publicly available data was lacking for cervical and ovarian tumours, we generated genome-wide methylome data for three high-grade serous ovarian carcinomas (HGSOC), two cervical adenocarcinomas and three cervical squamocellular carcinoma in-house and included it in the atlas. For interpretation purposes, we combined deconvoluted percentages for the two cervical cancer subtypes and from the colon and the rectal adenocarcinoma to identify the TOO.

Differentially methylated sites were selected by comparing the CpG site methylation ratio of one tissue against the rest of the entities in the reference (one-versus-all strategy) and then extended to regions. To ensure that the methylation marker regions were unique, a methylated or unmethylated region should have a distinct methylation pattern in one tissue versus the other entities in the reference atlas (Figure 1A). We identified 17874 differentially methylated regions across the genome for the 13 reference entities. The number of marker regions per reference entity ranged from 23 to 7058 with a median count of 5 CpGs and median length of 512bp (Figure 1B).

**Figure 1.**
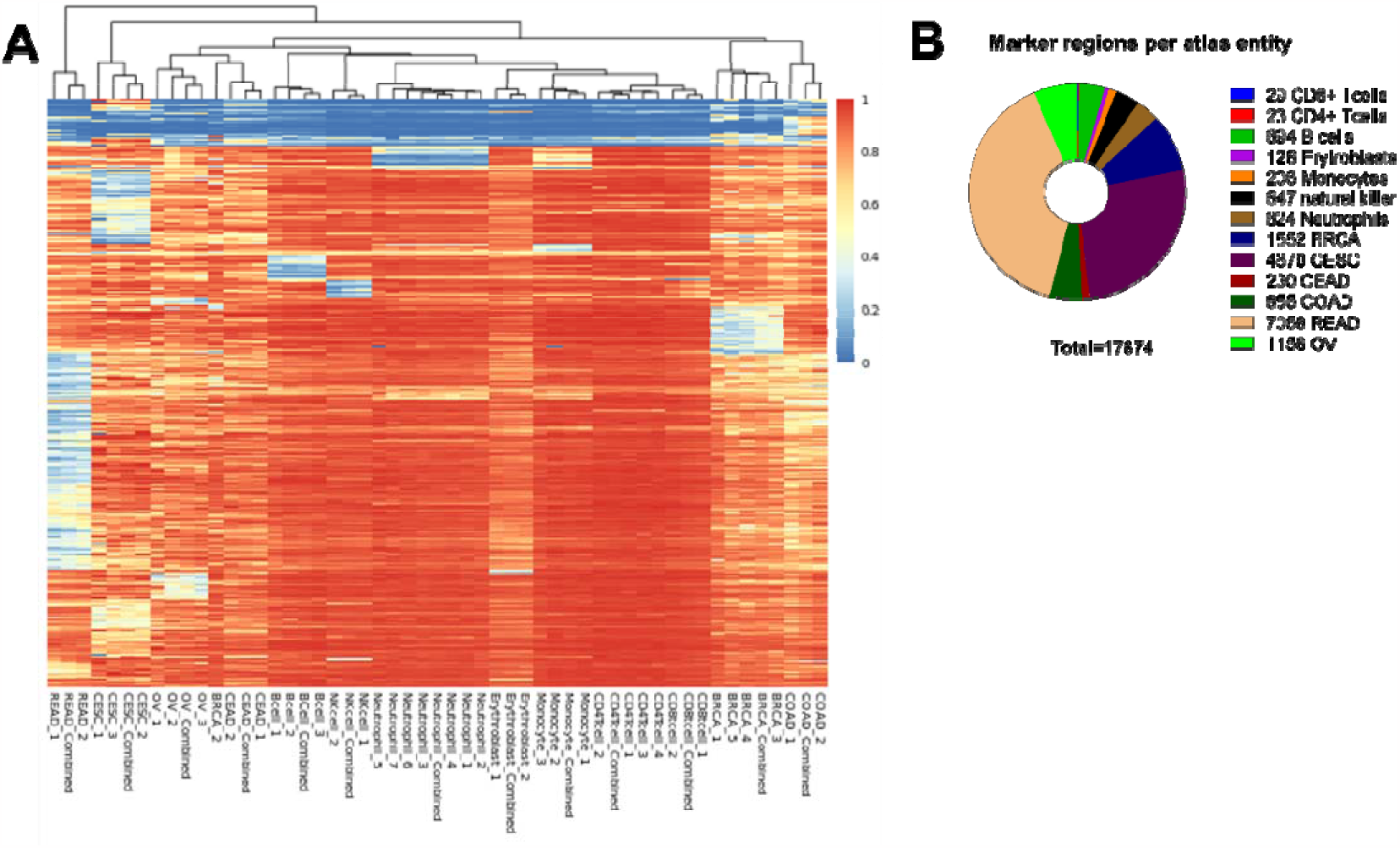
(A) Heatmap displaying the methylation ratio of the selected marker regions across the atlas entities. The methylation ratio is represented on the colour scale with red indicating a value close to one, meaning hypermethylation, and blue to zero, meaning hypomethylation. The individual samples per tissue/cell-type and the combined entity (named Combined, as described Materials and Methods) considered for the marker selection are shown on the X-axis. (B) Number of marker regions per atlas entity. BRCA, breast carcinoma; CEAD, cervical adenocarcinoma; CESC, cervical squamocellular carcinoma, COAD, colorectal adenocarcinoma; OVCA, ovarian carcinoma; READ, rectal adenocarcinoma.

### MetDecode is robust against noise and the presence of unknown contributors

Along with cancerous cells, cfDNA samples from cancer patients also contain other cell types, such as immune cells. A reference atlas of tumours and immune cells will often be incomplete and hence the analysis methods run the risk of being blinded for unknown contributions. While the primary goal of our method is to predict the type of cancer, standard supervised classifiers will fall short because (i) the number of training samples per class is too low (2 to 7 per class in this study) for standard classifiers, (ii) samples contain a variable proportion from cells from some classes, and (iii) samples might also contain cells of unknown types/profiles. Instead, we develop an algorithm to estimate the proportions from a mixture of reference samples also accounting for unknown cell-types. The method then assigns the sample to a particular tissue type by considering the atlas entity with the highest proportion falling outside the established normal range.

Contrary to methylation arrays or targeted sequencing which focus on specific loci, whole-genome methylation sequencing produces generally lower coverage due to the reads being spread over the entire genome. Therefore, the coverage of each CpG site is reduced and noise is exacerbated [23]. Not only does the genome coverage vary from sample to sample due to differences in sequencing depth, but it also varies along the genome itself (e.g., due to differences in mappability). Accordingly, we devised MetDecode to account for the reliability of the methylation ratio estimates (e.g., variability at methylation loci both in the atlas and cfDNA samples [35]) in the presence of noise. Since higher coverage of a marker region enables a more accurate estimate of its methylation ratio (under the assumption of the absence of biases, which we implicitly assumed), we re-weighted our objective function (see Methods) to lower the contribution of lower-coverage marker regions to the objective function.

The importance attached to the coverage is controlled by a hyper-parameter β, which determines the rate at which the weight of a marker region increases with its coverage. To assess the relevance of this new feature, we compared our method in default settings (*β* = 1) to our method without considering the coverage (*β* = 0). For this purpose, we designed simulations based on real data (see Supplementary Figure 1, Additional File 1), with random noise injection based on binomial distributions and deconvoluted these random mixtures. In Figure 2A, we reported the distribution of Pearson correlation coefficients between estimated and expected cell type proportions across 30 runs. When averaging the correlation coefficients across cell types (bottom right violin plot in Figure 2A), Pearson coefficient appears to be significantly higher in the (*β* = 1 setting (p<0.001; T-test; one-sided), highlighting the gain in deconvolution performance obtained when increasing the attention of our deconvolution algorithms on high-coverage regions, both in the atlas and cfDNA samples.

**Figure 2.**
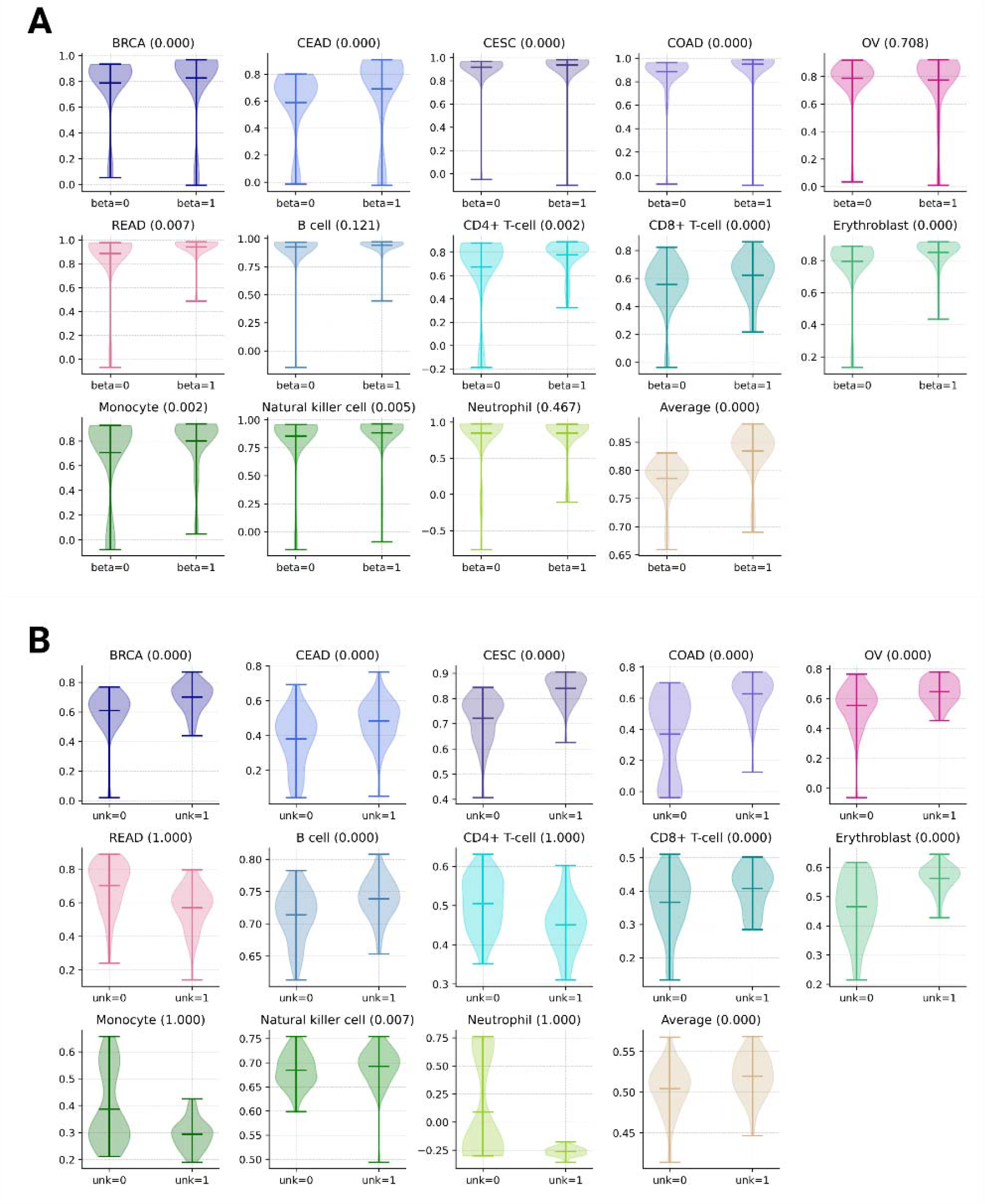
(A) Pearson correlation coefficients of MetDecode without () and with () consideration to the coverage across 30 simulation runs. For each cell type, a one-sided T-test has been performed to assess the difference in the distributions of Pearson coefficients, and the corresponding p-value reported between brackets. (B) Pearson correlation coefficients without (unk=0) and with exactly one (unk=1) unknown modelled by MetDecode. BRCA, breast carcinoma; CEAD, cervical adenocarcinoma; CESC, cervical squamocellular carcinoma, COAD, colorectal adenocarcinoma; OVCA, ovarian carcinoma; READ, rectal adenocarcinoma.

To create the atlas, only a limited number of cell and tissue types were selected. However, the cfDNA is made up of DNA derived from many different cell and tissue types, albeit usually in lower amounts. To account for this incompleteness of the atlas, we included the possibility to model unknown cell types. We opted for a data-driven approach that infers the unknowns using the cfDNA samples as well as the (incomplete) atlas, based on the residuals obtained after deconvolution (difference between the original and reconstructed/convoluted cfDNA samples). To demonstrate the relevance of this novel feature, we performed experiments analogous to those described above to quantify the performance characteristics of MetDecode when modelling one unknown cell type (unk=1), compared to the situation where the atlas is assumed to be complete (unk=0). In our simulations, cfDNA mixtures were defined as random linear combinations of the atlas entities (with proportions sampled from a Dirichlet distribution, see Additional File 1), plus an unknown entity with random binary methylation pattern. We observed a significant improvement of Pearson correlation coefficients across 30 runs (p<0.001; T-test; one-sided Figure 2B) for 5 out of the 6 cancer tissues included in our atlas. Overall, unknown modelling enhanced deconvolution accuracy for 9 out of the 13 cell types, and decreased performance for the 4 remaining cell types. When averaging the Pearson correlation coefficients across all the cell types (bottom right violin plot in Figure 2B), we observed a p-value of 0.001. These results highlight the relevance of unknown modelling when unknown cell types in the sample of interest have their methylation patterns uncorrelated with the atlas entities.

### MetDecode identifies the correct TOO in genomic DNA from leukocytes and tumour tissue

To evaluate the accuracy of MetDecode for deconvoluting and assigning the correct tissue type, we applied the method on 12 WBC-derived gDNA methylomes from healthy controls (mean age: 48.08 years; range: 22-77; M/F:5/7) and 20 gDNA methylomes from tumour tissue biopsy of breast (n=5), colorectal (n=6), cervical (n=6) and ovarian (n=3) cancer (Figure 3).

**Figure 3.**
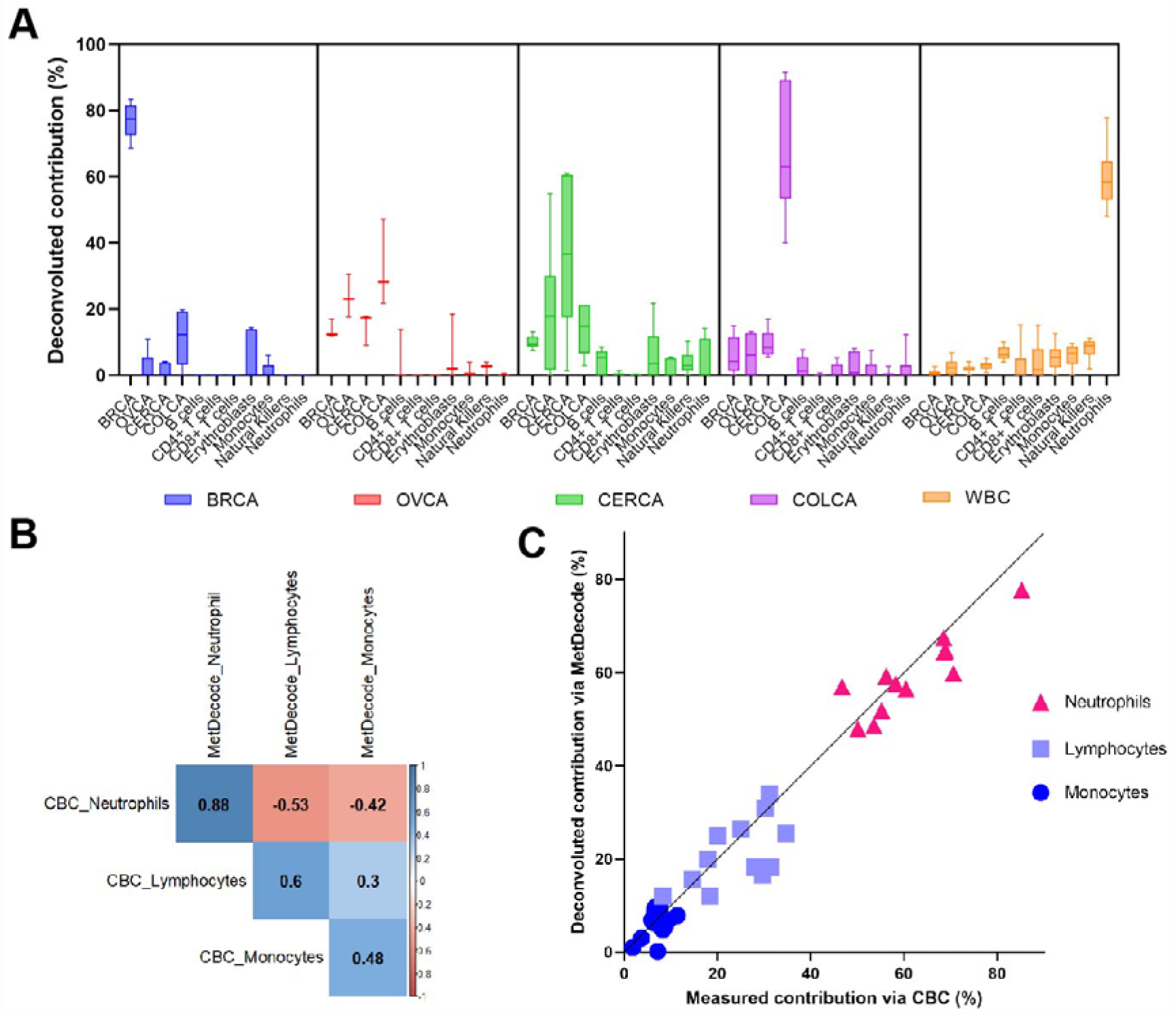
Deconvolution of genomic DNA methylation profiles using MetDecode. (A) Deconvolution of gDNA derived from WBC and tumour FFPE biopsies. For each sample group, the deconvoluted contribution from every atlas entity is shown. The box represents the interquartile range, the extremity represents the minimum and the maximum value. The median is marked with a horizontal bar inside the box. WBC, white blood cells; BRCA, breast carcinoma; CERCA, cervical carcinoma; COLCA, colorectal carcinoma; OVCA, ovarian carcinoma. (B) Pearson correlation between deconvoluted contributions measured via complete blood count (CBC) and MetDecode for neutrophils, monocytes and lymphocytes. (C) Relative contribution measured via CBC (X-axis) vs the deconvoluted proportion estimated using MetDecode (Y-axis) for the neutrophils, lymphocytes and monocytes in the gDNA derived from WBC. The diagonal line represents the identity line.

When deconvoluting, the methylomes are distributed amongst different atlas entities. When the major contributor amongst all the atlas entities was the expected tissue, the assignment was considered correct. The healthy controls were considered as correctly assigned when neutrophils were deconvoluted as the main contributor [36]. MetDecode assigned the correct tissue in 29 out of 32 samples (Overall Accuracy: 90.63%). All WBC-derived samples showed neutrophils as the main contributor. All 5 breast tumour samples and 6 colorectal samples were assigned to their respective cancer. In addition, 5/6 and 1/3 cervical and ovarian tumours were classified correctly. One of the 6 cervical samples was classified as an ovarian tumour. In addition, two out of 3 ovarian tumour samples (n=2 clear cell carcinoma) were misclassified as colorectal cancers.

To assess the accuracy of mixture deconvolution, we compared the results of the WBC deconvolution to Complete Blood Counting (CBC) using matched blood samples. We observed a high correlation for the neutrophils fraction (r=0.879, p-value<0.001, Figure 3B), when comparing CBC and MetDecode deconvolution estimates. Lower correlation was found for lymphocytes and monocytes (r=0.60; r=0.48, respectively). However, this is similar to other reports [34,37]. In conclusion, these results demonstrate that MetDecode can identify major contributors in samples containing mixture of cell-types.

### Metdecode allows accurate deconvolution of in-silico mixes

We next evaluated the limit of detection and assessed the concordance in the relative contribution estimation of MetDecode using *in-silico* mixtures of tumour DNA. Breast, colorectal, cervical and ovarian cancer data was combined with cfDNA from healthy donors at ratios varying from 50-0.1% to reflect an average depth of 6X (see Additional File 1). Deconvolution of the *in-silico* mixes showed that MetDecode detects tumour DNA proportions down to 4.3%. The mean correlation between the expected and the deconvoluted percentage was r=0.8603 (p<0.001). The highest correlation was obtained for the breast cancer *in-silico* mixes, followed by ovarian, cervical and colorectal (0.998, p<0.001; 0.982, p<0.001; 0.976, p<0.001 and, 0.866 p<0.001, respectively, Figure 4). The expected and estimated percentages of the spiked-in component show a strong correlation, indicating that MetDecode’s relative proportion estimations are indeed reliable.

**Figure 4.**
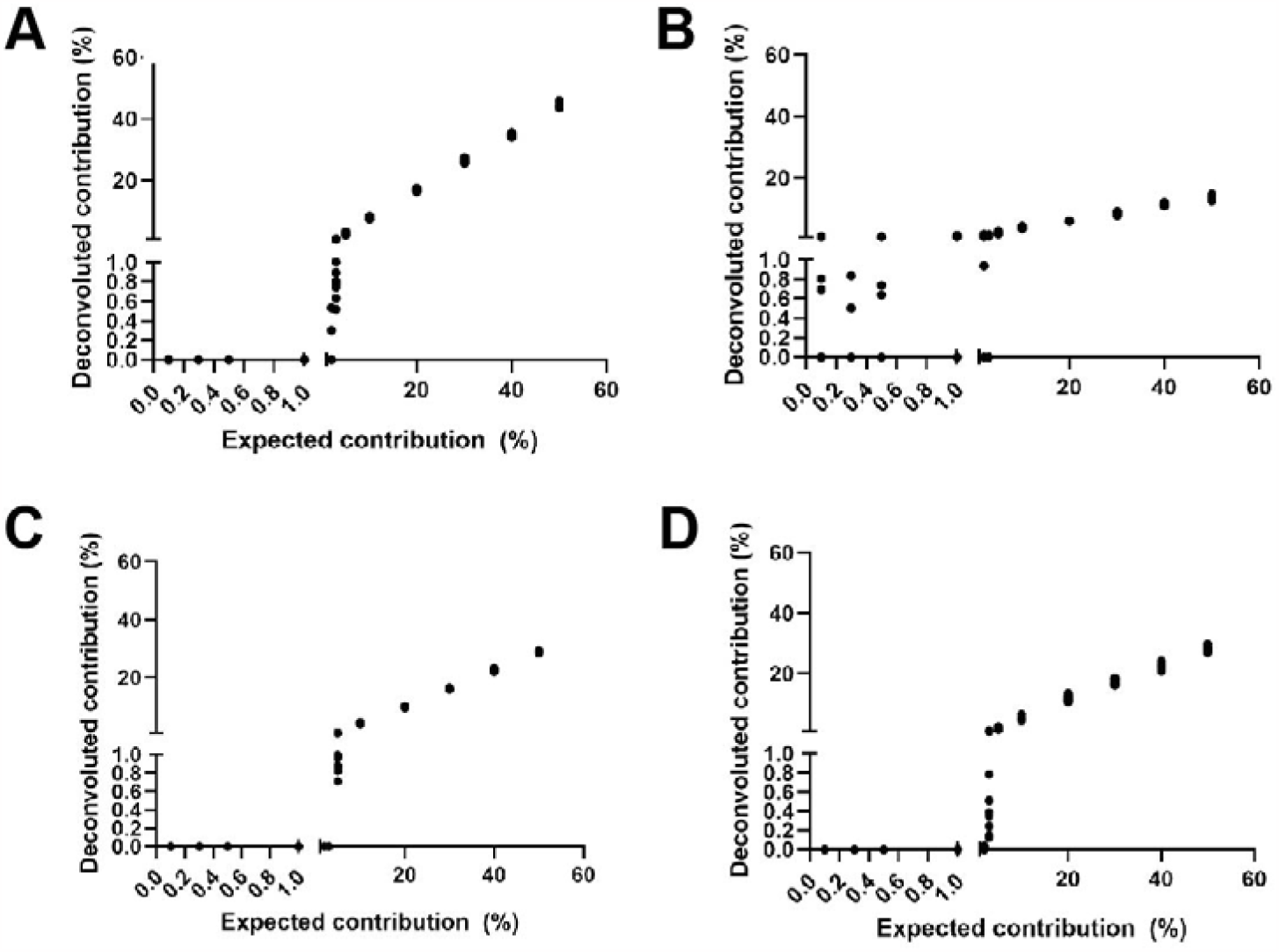
Correlation plots between deconvoluted and expected contribution in percentage of the tumour tissue spiked-in in the *in-silico* mixes. Random reads were combined from a healthy control BAM file and a tumour gDNA BAM file to create an *in-silico* mix and repeated 10 times to obtain replicates (A), (B), (C), (D) represent the *in-silico* mixes for breast, ovarian, colorectal and cervical cancer, respectively. Each dot represents the value for a replicate with a deconvoluted % (y-axis) vs expected % (x-axis) of tumour tissue DNA.

### MetDecode correctly identified the tumour origin in cfDNA from cancer patients

MetDecode was subsequently applied to whole-genome cfDNA methylation sequencing data from healthy controls (n=93; mean age: 66.8 years; range:18-90) and treatment-naive patients with a confirmed cancer diagnosis (n=16; n=5 breast, n=4 colorectal, and n=7 ovarian cancers; Table 1). We selected samples with a minimum tumour fraction (TF) of 3% (measurement based on ichorCNA [31]).

**Table 1.** Clinical information of the cancer cohort and deconvolution outcome.

| Sample | Sex | Age | Cancer type | Cancer subtype | Stage | TOO assigned | MetDecode (%)<sup>*</sup> | TF (%) |
| --- | --- | --- | --- | --- | --- | --- | --- | --- |
| Case 1 | F | 80-89 | Breast cancer | Triple Negative | IIIA | BRCA | 3.398 | 3.96 |
| Case 2 | F | 50-59 | Breast cancer | Luminal B | IIIA | BRCA | 5.02 | 5.86 |
| Case 3 | F | 40-49 | Breast cancer | Triple Negative | IA | **NA** | **NA** | 4.44 |
| Case 4 | F | 50-59 | Breast cancer | Her2 positive | IV | BRCA | 14.014 | 6.27 |
| Case 5 | F | 50-59 | Breast cancer | Triple Negative | IV | **COLCA** | 12.439 | 13.66 |
| Case 6 | F | 80-89 | Colorectal cancer | Unknown | IIIC | COLCA | 33.909 | 10.89 |
| Case 7 | M | 70-79 | Colorectal cancer | Unknown | IVA | COLCA | 40.852 | 15.14 |
| Case 8 | M | 80-89 | Colorectal cancer | MSS | II | COLCA | 13.693 | 3.36 |
| Case 9 | M | 60-69 | Colorectal cancer | MSS | IV | COLCA | 21.473 | 11.38 |
| Case 10 | F | 50-59 | Ovarian cancer | Mucinous carcinoma | IVB | **CERCA** | 5.793 | 12.25 |
| Case 11 | F | 70-79 | Ovarian cancer | LGSOC | IIIC | OVCA | 20.682 | 19.07 |
| Case 12 | F | 60-69 | Ovarian cancer | Endometrioid carcinoma | IA | OVCA | 13.388 | 7.91 |
| Case 13 | F | 40-49 | Ovarian cancer | LGSOC | IIIC | OVCA | 11.039 | 5.50 |
| Case 14 | F | 60-69 | Ovarian cancer | HGSOC | IIIC | OVCA | 22.998 | 11.14 |
| Case 15 | F | 60-69 | Ovarian cancer | HGSOC | IIIA | OVCA | 9.439 | 4.47 |
| Case 16 | F | 50-59 | Ovarian cancer | HGSOC | IV | OVCA | 49.481 | 27.12 |

<sup>\*</sup> The number reported refers to the deconvoluted percentage of the putative TOO.
*In bold are indicated the misassigned TOO.*
TF, Tumour Fraction; TOO, Tissue Of Origin; MSS, Microsatellite Stable; LGSOC, Low-Grade Serous Carcinoma; HGSOC, High-Grade Serous Carcinoma; BRCA, breast carcinoma; CERCA, cervical carcinoma; COLCA, colorectal carcinoma; OVCA, ovarian carcinoma; NA, not assigned.

cfDNA data from the healthy individuals was used as control to establish the reference range of each atlas entity (Supplementary Table 2, Additional File 2). As expected, [22,34,38] neutrophils were the major contributors to plasma cfDNA (37.51%±9.418), followed by erythroblasts (19.34%±4.625), and monocytes (18.16%±3.908) (Supplementary Figure 3, Additional File 1).

We then investigated the deconvoluted composition of the cancer patient-derived plasma cfDNA and compared it to the normal ranges in healthy controls. Overall, the tissue in which the primary tumour resides was increased in all three cancer types. The colorectal cancers showed an average 35.67-fold increase in COLCA contribution compared to the healthy controls, ovarian cancers an average 8.06-fold increase in OVCA contribution and breast cancers an average 7.8-fold increase in BRCA contribution (Figure 5A). This shows that tissue-specific signals are well classified.

**Figure 5.**
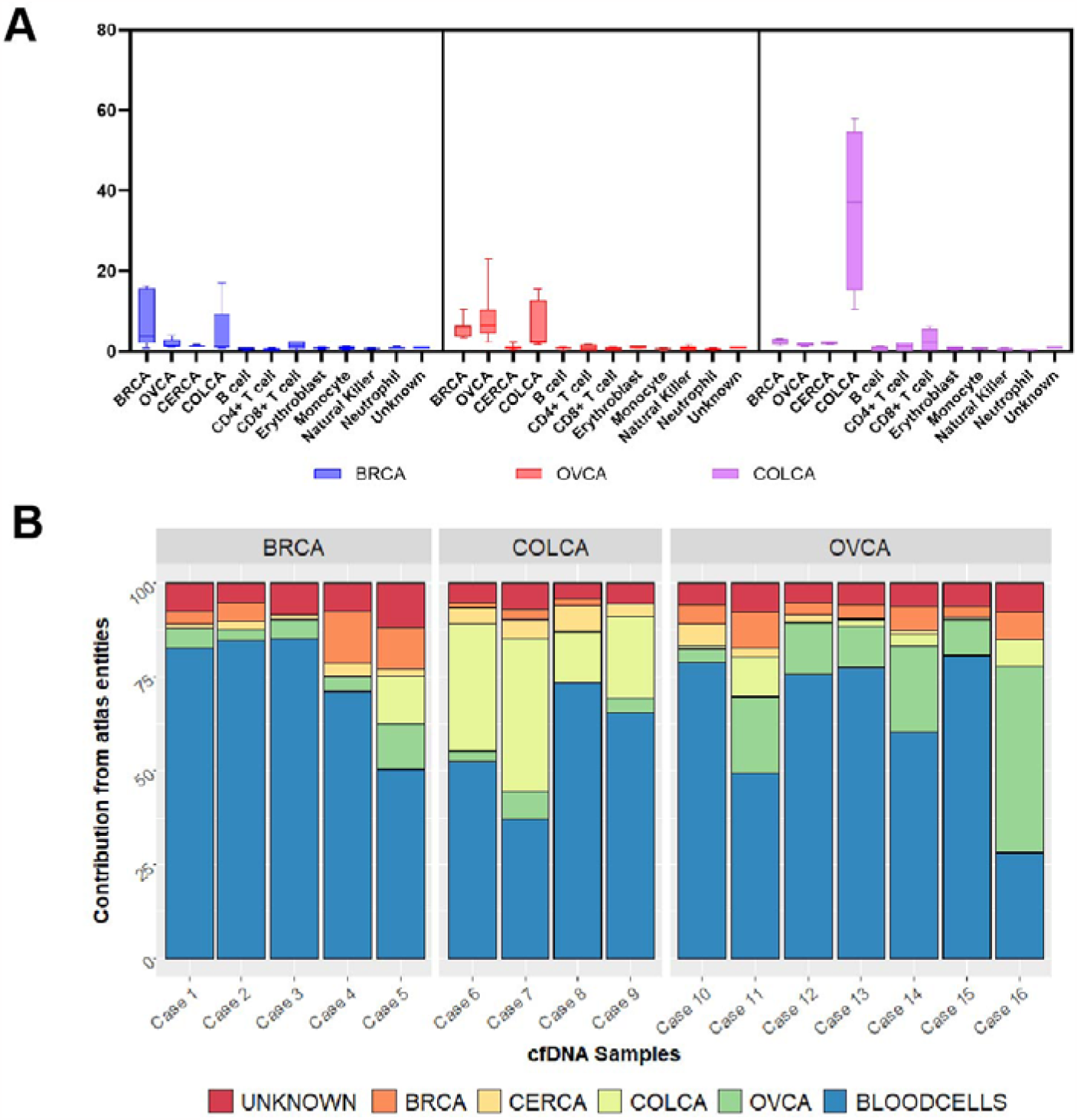
Deconvolution of the cfDNA derived from patients with a confirmed cancer diagnosis. (A) Fold enrichment of the deconvoluted percentages in the cfDNA from each cancer cohort was determined using healthy controls as the baseline. For each group of the cohort, namely breast, ovary and colorectal, the deconvoluted contribution of each atlas entity is represented in the box plot. The box represents the interquartile range, the extremity represents the minimum and the maximum value. The median is marked by a horizontal bar inside the box. (B) Distribution of the deconvoluted percentage in the 16 cfDNA samples from cancer patients. The contribution from the different blood cell types is summed up and shown in blue. Expected range for each atlas entity was established using mean±2SD of the contribution detected in healthy controls. An atlas entity is referred to as the tissue of origin (TOO), when the relative contribution in that specific tissue is higher than this expected range. If multiple entities fall outside the range, the highest is considered the putative TOO. BRCA, breast carcinoma; CERCA, cervical carcinoma; COLCA, colorectal carcinoma; OVCA, ovarian carcinoma

We next assessed the ability of MetDecode to assign the correct TOO in these cfDNA samples from cancer patients. Among the deconvoluted values falling outside the normal range established, the highest contributor across the cancer components of the reference atlas was regarded as the putative TOO of the malignancy. Overall, MetDecode assigned the correct TOO in 13 out of 16 cancer cases (accuracy 81.25%). 100% of the colorectal and 85.71% of the ovarian cancer cfDNA samples were correctly classified. One mucinous ovarian carcinoma (stage IV) was predicted to have cervical cancer tissue as a major cancer contributor. For breast cancer, 60% of the samples were assigned to the correct tissue. One triple-negative breast cancer sample (case 5, stage IV) was misclassified as colorectal. For this sample, the deconvoluted contribution from several atlas entities, namely breast, ovary and the unknown component were also higher than normal (Supplementary Table 3, Additional File 2). This result might be caused by metastasis in multiple organs, such as liver, lung and lymph nodes. Additionally, one triple-negative breast cancer cfDNA sample (case 3, stage I) did not show any alteration compared to the controls.

## Discussion

While there are huge efforts to enable cfDNA-based early multi-cancer detection, the utility of such tests is being explored [39]. Here, we present a novel method for methylation-based cfDNA deconvolution to aid the identification of cancer types. MetDecode combines a methylome reference with a novel algorithm which can model for unknown contributors absent in the reference and is mindful of the coverage of each marker in the reference. In addition to accurately estimating the tumour proportion in in-silico mixtures (r=0.8603, p<0.001), the method also assigned the correct TOO in 81.25% of the cfDNA samples from cancer patients.

Other deconvolution methods using epigenetic markers have been developed. MethAtlas [22] and cfNOMe [28], built on non-negative least squares and constrained programming respectively, modelled cfDNA mixtures as perfect linear combinations of the reference cell types. cfNOMe performed a multimodal analysis by complementing methylation with nucleosome occupancy profiles. To account for incomplete reference atlases, CelFiE [23] extended previous approaches by inferring the methylation patterns of unknown cell types from the data directly using a probabilistic model. MethylCIBERSORT [40] built on support vector regression (SVR) to perform robust deconvolution and discard the effect of markers with low reconstruction error whereas ARIC [24], also based on SVR, introduced a feature selection step to remove redundant markers, using condition numbers as a measure of collinearity. MethylResolver [25], on the other hand, alleviated the effect of outliers by using a least trimmed squares approach. CancerLocator [20] used a probabilistic model to estimate the tumour burden and identify the correct cancer type. CancerDetector [21] achieved higher sensitivity than CancerLocator by performing cancer classification at the level of individual sequencing reads. However, neither of these two methods allows full deconvolution of white blood cells but rather estimates the cancer proportions alone, therefore limiting the interpretability of the results. Finally, MeDeCom [26] is a reference-free approach based on regularized matrix factorization.

Among all these methods, CelFiE is the first and only full reference-based technique proposing to tackle issues related to both the non-completeness of the atlas and the noisy nature of sequencing data. However, the number of parameters in CelFiE’s underlying model scales to the number of markers and (unknown) cell types, therefore exposing the method to overfitting risks. Full tissue–type deconvolution with modelling of unknowns has, to our knowledge, only been proposed in CelFiE and has not been properly addressed in the literature, as described above. Here we propose a novel computational method, coined MetDecode, to disentangle the methylation patterns of different cell types contributing to cfDNA mixtures, while accounting for the potential incompleteness of the reference atlas and the inaccurate estimation of methylation ratios in low-coverage regions.

Incorporating these algorithms into our pipeline was not practically feasible, since the peculiarities of our data make them mostly unsuitable for most of these deconvolution approaches. For example, MeDeCom [26] was not designed to handle a reference atlas, as the method is unsupervised. MethAtlas [22], like many other methods, does not account for the coverage, as the method expects methylation array data as input instead of sequencing data. Finally, CfNOMe [28] requires nucleosome occupancy profiles which we did not compute, as such profiles are not handled by some of the other tools (e.g. CelFIE [23]).

The deconvolution of in-silico mixes affirms the performance of our deconvolution method. With respect to the gDNA samples, 2 ovarian carcinomas were misclassified as colorectal cancer. We hypothesize that the misassignment results from not including different ovarian carcinoma subtypes to build the reference atlas. In fact, the subtype used to build the reference atlas was high-grade ovarian carcinoma, while the three gDNA test samples were classified as clear cell ovarium carcinoma (n=2) and mucinous carcinoma (n=1). Similar to what was observed in the deconvolution of gDNA, one cfDNA sample from an ovarian cancer patient was not correctly classified. We hypothesize that the misassignment is a consequence of the absence of these ovarian carcinoma subtypes in the reference atlas and hence remains unrecognised [41,42].

A way to overcome this limitation might reside in using cell-type specific methylation data for the reference atlas creation [38]. The development of cell-type-based methylome atlases might provide an opportunity to dissect the different contributing cell types and may well outperform the methylome markers based on bulk tissue-specific entities. Cell-type specific methylation would ensure more precise deconvolution and offer clearer insight into the origin of the tumour. However, cell-type-based methylation data is not yet available for the cancer tissues of interest [22]. Similarly, including different subtypes for the marker selection could potentially allow subtype identification and improve overall cancer diagnosis. Given the small number of samples available, no clear correlation between (mis)classification and cancer stage could be drawn.

Unique to our approach of selecting methylation markers is that we select regions with a methylation pattern distinct to one cell or tissue type and depending on the aims of the end user, it can be applied to different tissues or cell types. Existing marker selection approaches seek to maximise the difference between methylation ratio and the median of the ratio across all tissues. This does not necessarily ensure that only one tissue or cell type is differentially methylated compared to the rest of the entities in the reference atlas [23]. The limitation of our approach is that the number of markers may reduce with an increment in the number of atlas entities. Additionally, our methylome atlas was built with a limited number of samples per atlas entity. We envision that increasing the number of samples per atlas entity may improve specificity of the selected methylation markers.

## Conclusions

Deconvolution of the cfDNA epigenetic signatures is an elegant approach to deduce the TOO or cancer-type. To estimate the contributions and the type of cancer and white blood cell types in a cfDNA sample, we developed MetDecode, a methylome reference-based deconvolution algorithm. MetDecode can model the unknown contributors unavailable in the reference and account for the coverage of each marker region to alleviate the potential sources of noise.

Despite the limited sample size, the results presented here are encouraging and important for the future of liquid biopsy clinical application. In fact, a tool able to pinpoint the TOO of a malignancy can be used to streamline the diagnostic process in cancer patients. Emblematic cases in which the TOO detection via cfDNA can be of clinical utility are in the detection of cancer-like signals in maternal blood during routine non-invasive prenatal screening and in case of metastatic tumours of unknown primary. Deconvoluting and defining the TOO will aid the oncologists in identifying the tumour and direct treatment, streamlining the diagnostic process, especially in cases in which invasive examinations and radiological investigation are not ideal. Furthermore, if specific immune characteristics of the malignancy could be detected thanks to the blood-derived entities residing in the atlas, an important dowel for the treatment decision of the patient can be added simultaneously to the TOO identification. To conclude, we developed a method for deconvoluting the components of plasma enabling detection of cancer origin using tissue-specific methylome information.

## Data Availability

The data that support the findings of this study is in controlled access data storage in EGA under EGAS00001007493 and is available upon reasonable request. MetDecode is available on GitHub as a Python package: https://github.com/AntoinePassemiers/MetDecode

## List of abbreviations

cfDNA: cell-free DNA
TOO: tissue of origin
CNA: copy number alterations
FFPE: formalin-fixed paraffin-embedded
WGBS: whole genome bisulfite sequencing
NNLS: non-negative least squares
MSE: mean squared error
HGSOC: high-grade serous ovarian carcinoma
WBC: white blood cells
CBC: complete blood counting
TF: tumour fraction
MSS: microsatellite stable
LGSOC: low-grade serous carcinoma
BRCA: breast carcinoma
CERCA: cervical carcinoma
COLCA: colorectal carcinoma
OVCA: ovarian carcinoma
NA: not assigned
SVR: support vector regression

## Declarations

### Ethics approval and consent to participate

The study was approved by the ethical committee of the University Hospitals Leuven (study protocols S62285, S62795, S63983, S66450, S59207 and S51375).

### Consent for publication

Not applicable

## Authors contribution

D. Sudhakaran, S. Tuveri, T. Jatsenko, L. Lenaerts, and J.R. Vermeesch conceptualized and designed the study. S. Tuveri, T. Jatsenko, L. Lenaerts, Laga T., Punie K., Tejpar S., Coosemans A., Testa A., Ficherova D., Nieuwenhuysen E, Timmerman D. and Amant F. carried out clinical sample collection. G. Floris, A.S. Van Rompuy and Sagaert X. performed diagnosis on the tissue and selected the paraffin material to be used in the study. S. Tuveri performed the wet-lab tasks and coordinated sequencing of cfDNA and gDNA. D. Sudhakaran designed the marker selection and performed bioinformatics analysis. A. Passemiers, D. Raimondi and Y. Moreau conceptualized the algorithm. A. Passemiers designed and implemented the algorithm. S. Tuveri, T. Jatsenko, L. Lenaerts, and D. Sudhakaran contributed to the interpretation of results. S. Tuveri, D. Sudhakaran, and A. Passemiers wrote the manuscript; all co-authors reviewed the manuscript.

## Acknowledgements

We would like to thank the patients and healthy blood donors for their availability and Dr Leen Vancoillie and Dr Ilse Parijs for their help in extracting cfDNA from plasma samples.

