## Supplemental file 1 for "MetDecode: methylation-based deconvolution of cell-free DNA for non-invasive multi-cancer typing"

**Additional material**

***Deconvolution algorithm***

*Overview, notation and modelling*

A reference methylation atlas is provided in the form of two matrices $M^{\left( atlas \right)} \in\mathbb{N}^{m\times p}$and $D^{\left( atlas \right)} \in\mathbb{N}^{m\times p}$, in which $D_{jk}^{\left( atlas \right)}$ is the number of CpG sites spanned by reads in the marker region $r$ for reference cell type $j$ and $M_{jk}^{\left( atlas \right)}$ is the number of methylated ones. Similarly, cfDNA data is supplied as two matrices $M^{\left( cfdna \right)} \in\mathbb{N}^{n\times p}$ and $D^{\left( cfdna \right)} \in\left[ 0, 1 \right]^{n\times p}$, where $n$ is the number of patients. Because each CpG site can be spanned by more than one sequencing read, in practice each site contributes to the counts more than once.

The methylation ratios of the atlas and the cfDNA samples are computed as $R_{jk}^{\left( atlas \right)} = \frac{M_{jk}^{\left( atlas \right)}}{D_{jk}^{\left( atlas \right)}}$ and $R_{ik}^{\left( cfdna \right)} = \frac{M_{ik}^{\left( cfdna \right)}}{D_{ik}^{\left( cfdna \right)}}$, respectively. The goal of our algorithm is to estimate a matrix $A \in\left[ 0, 1 \right]^{n\times m}$, where $A_{ij}$ is the estimated proportion of cell type $j$in cfDNA profile $i$. Before inference, we add $h$ extra rows to matrix $R^{\left( atlas \right)}$ to model the presence of unknown cell types potentially present in the cfDNA mixtures. $h$ is a hyper-parameter that can be tuned by the end-user.

*Modelling of unknown contributors*

We first compute the lower bounds $R_{k}^{\left( lb \right)} = \min_{j} R_{jk}^{\left( atlas \right)}$ and upper bounds $R_{k}^{\left( ub \right)} = \max_{j} R_{jk}^{\left( atlas \right)}$ on the atlas values. The initial cell type proportion estimates $\alpha$are obtained by the non-negative least squares (NNLS) algorithm. We quantify the excess of methylation of the reconstructed samples in marker region $k$ as the median residual $e_{k} = median_{i} \left( -R_{ik}^{\left( cfdna \right)} + \sum_{j} \alpha_{ij}R_{jk}^{\left( atlas \right)} \right)$. Intuitively, $e_{k}$ is strictly greater (smaller) than 0 when most of the reconstructed cfDNA samples produced methylation ratios higher (lower) than what can be observed in $R^{\left( cfdna \right)}$. Therefore, $e_{k}$ provides a hint on the information currently lacking from the atlas. The new row of $R^{\left( atlas \right)}$ is defined as follows: its $k$th element will be set to $R_{k}^{\left( lb \right)}$ when $e_{k}$ is positive, and vice versa. Corresponding row in $D^{\left( atlas \right)}$ is simply computed as the median read counts across the samples. Corresponding row in $M^{\left( atlas \right)}$ is determined by the element-wise product of methylation ratios and read counts. Once an extra row has been added to $D^{\left( atlas \right)}$, $M^{\left( atlas \right)}$ and $R^{\left( atlas \right)}$, the whole procedure is repeated until the desired number of unknown contributors is reached.

*Objective function*

We define the reconstruction error as the weighted average squared error between the original matrix of methylation ratios $M^{\left( cfdna \right)}$ and the reconstructed matrix. The weights are given by $W_{ik}^{\left( cfdna \right)} = \frac{\left( D_{ik}^{\left( cfdna \right)} \right)^{\beta}}{\sum_{a=1}^{n} \sum_{l=1}^{p} \left( D_{al}^{\left( cfdna \right)} \right)^{\beta}}$, where $\beta$ controls the importance of coverage in the deconvolution problem. The reconstruction error is formulated as $f\left( A \right) =\sum_{i=1}^{n} \sum_{k=1}^{p} W_{ik}^{\left( cfdna \right)}\left( M_{ik}^{\left( cfdna \right)} - \sum_{j=1}^{q} A_{ij}R_{jk}^{\left( atlas \right)} \right)^{2}$, and our deconvolution algorithm aims at minimizing this objective function by gradient-based optimisation.

*Inference*

Parameter $A$ is found by gradient-based optimization, using pytorch Python package (which ensures full and automated differentiation) and the Adam optimizer. The learning rate is decreased by a 0.9 factor when the update of the corresponding parameter resulted in an increase of the objective function at previous iteration and increased by a 1.015 factor otherwise.

Since the elements of $A$ are positive and its rows are summing up to one, $A$ is lying on a multinomial manifold. To ensure that $A$ remains on the manifold while optimizing, we instead optimize a matrix $A^{'}$ and apply a SoftMax function on the rows of $A^{'}$ to obtain a matrix $A$ with desired properties.

***Simulation procedure***

In Suppl. Fig. 1, we illustrate the data generation process used in our simulations. To simulate the fact that our atlas is not complete, we generated random methylation ratios using a $Bernoulli\left( 0.7 \right)$ distribution and appended each unknown cell type as a new row to $R^{\left( atlas \right)}$. The methylation ratios of cfDNA samples $R^{\left( cfdna \right)}$ have been generated as random linear combinations $\alpha$ of rows from $R^{\left( atlas \right)}$. Each row of $\alpha$ has been randomly sampled from a Dirichlet distribution with the following parameters for each cell type: Cancer=0.7393, B cell=3.0938, CD4=8.2576, CD8=3.8222, Erythroblast=1.7946, Monocyte=4.6937, Natural killer cell=2.6914, Neutrophil=29.6514, Unknown=15.000. The cancer proportion was randomly assigned to one cancer tissue at each run. To simulate the presence of noise, methylated counts have been randomly sampled from binomial distributions using the coverages from our original data. More specifically, the atlas values were defined by $M_{jk}^{\left( atlas \right)} \sim B\left( D_{jk}^{\left( atlas \right)}, R_{jk}^{\left( atlas \right)} \right)$ and cfDNA values by $M_{ik}^{\left( cfdna \right)} \sim B\left( D_{ik}^{\left( cfdna \right)}, R_{ik}^{\left( cfdna \right)} \right)$.

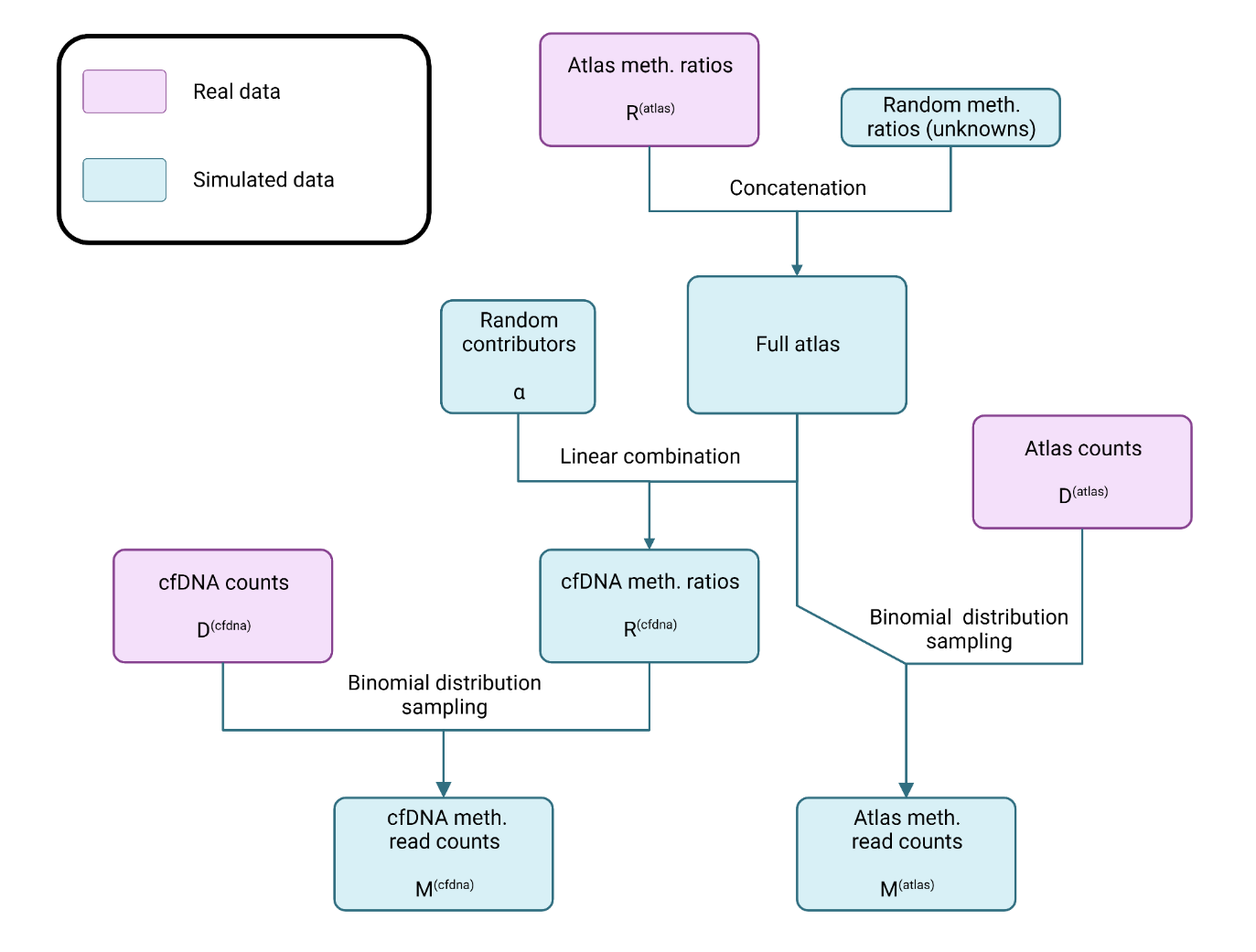
Supplementary figure 1: Flowchart of the simulation procedure. We used the counts and methylation ratios from our real atlas to generate the methylation ratios of random cfDNA mixtures, and used the cfDNA counts to determine the coverage of each marker region in these mixtures.

***Sample pre-processing for selecting methylation markers***

The downloaded data from all the public databases were converted to a uniform 2bp format so that every CpG site is represented in one line. Lift over to hg38 was done where required using in-house scripts. We observed high variance in the methylation ratio of few sites in samples of the same tissue/cell type (Supplementary figure 2). To avoid sample-specific bias, sites with a high variance (var>0.125) were removed. Multiple samples for one tissue/cell type were then combined by summing up the total reads and methylated reads at each CpG site to improve the coverage. This resulted in a “Combined” file with methylation ratio of CpG sites per reference atlas entity which was then used for selecting methylation markers. This process was also followed for the genomic samples processed in-house.

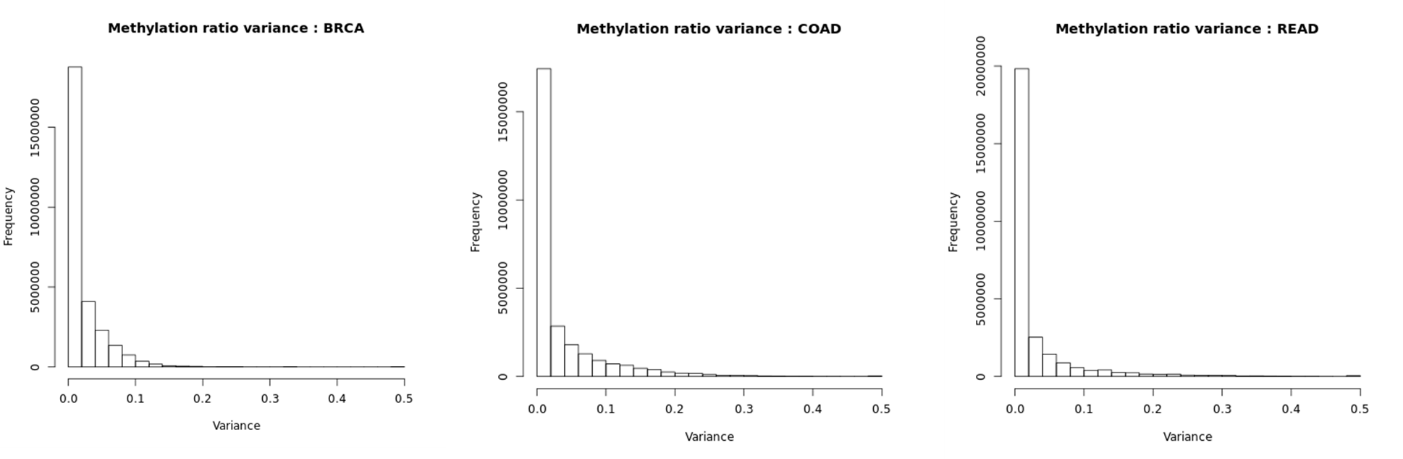
Supplementary figure 2: The distribution of variance of methylation ratio at every CpG site across samples from the same tissue type. Tumour samples downloaded from TCGA are shown here as an example. BRCA-Breast invasive carcinoma, COAD-Colon adenocarcinoma, READ-Rectum adenocarcinoma.

***Creation of in-silico mixtures***

*In-silico* mixtures were created using the aligned BAM files from a healthy control cfDNA and a tumour gDNA from a cancer type. The depth of an *in-silico* mix was set to 6X and corresponding read counts from a healthy control cfDNA and tumour gDNA BAM files were calculated based on the 12 tumour fractions (50, 40, 30, 20, 10, 5, 3, 2, 1, 0.5, 0.3 and 0.1). 10 replicates per tumour fraction (120 files in total) were generated by random selection of reads from the BAM file. The pairs of BAM files were then merged using samtools (v1.9) and methylation calling was performed using MethylDackel (v0.5.1). 2 samples from each cancer type namely breast, cervical, colorectal and ovarian were processed in the same manner resulting in 960 BAM files.

***Deconvolution of cfDNA from healthy patients***

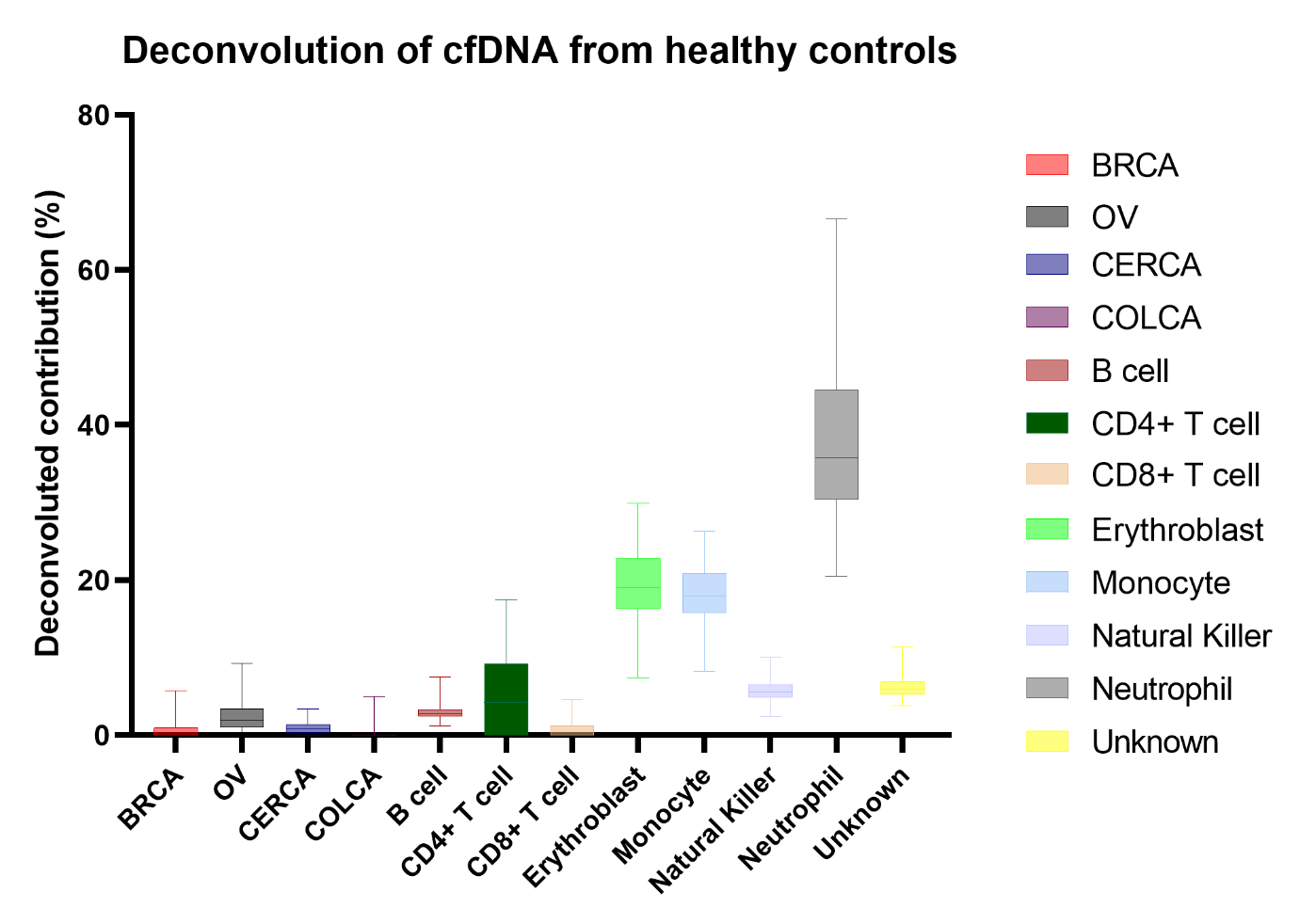

Supplementary figure 3. Deconvolution estimates for each atlas entity of the plasma cfDNA in 93 healthy individuals. The box represents the interquartile range, the extremity represents the minimum and the maximum value. The median is marked with a vertical bar inside the box. BRCA, breast carcinoma; CERCA, cervical carcinoma; COLCA, colorectal carcinoma; OVCA, ovarian carcinoma
