## Supplementary figures and images for "MetDecode: methylation-based deconvolution of cell-free DNA for non-invasive multi-cancer typing"

### Supplemental Figure 1

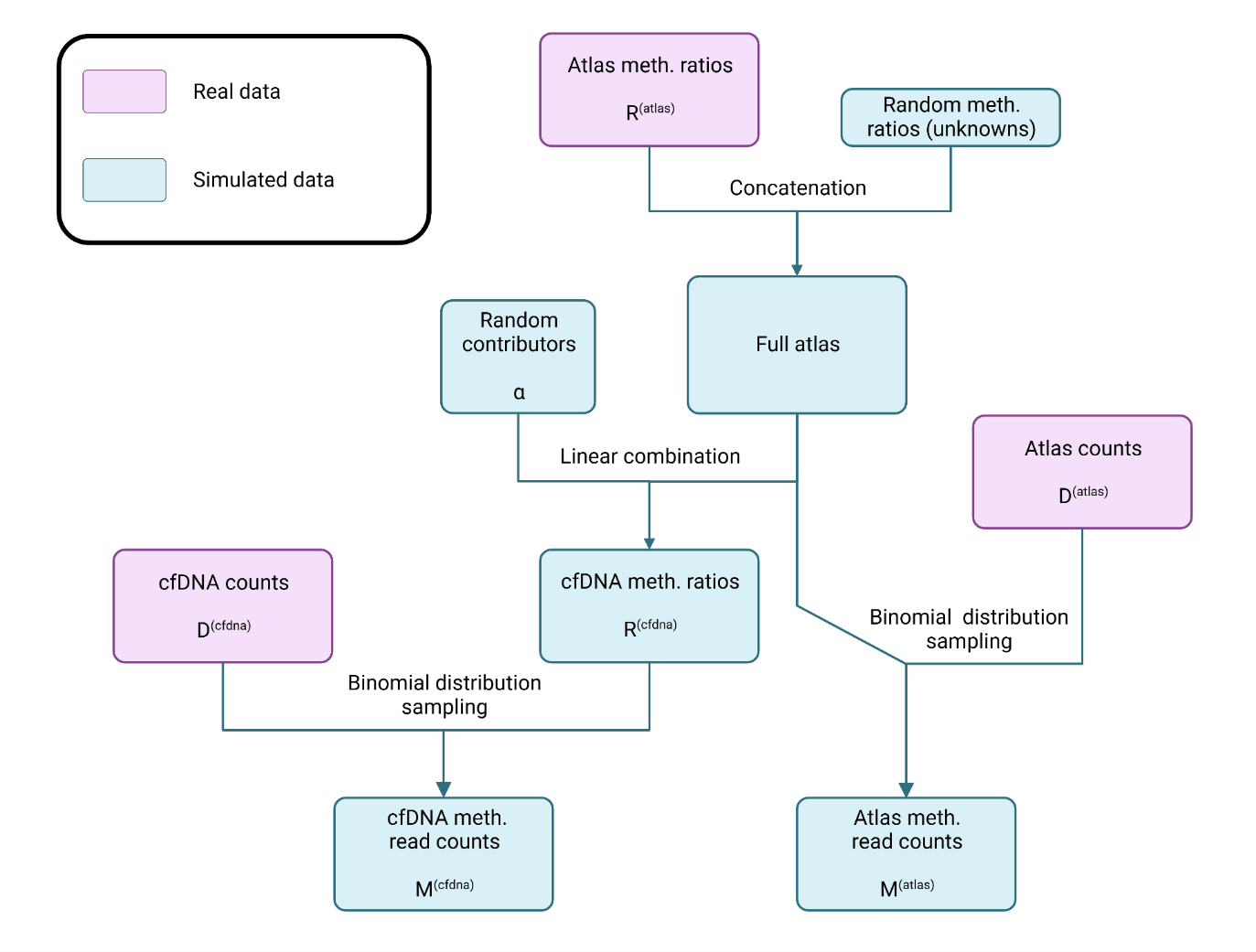

### Supplemental Figure 2

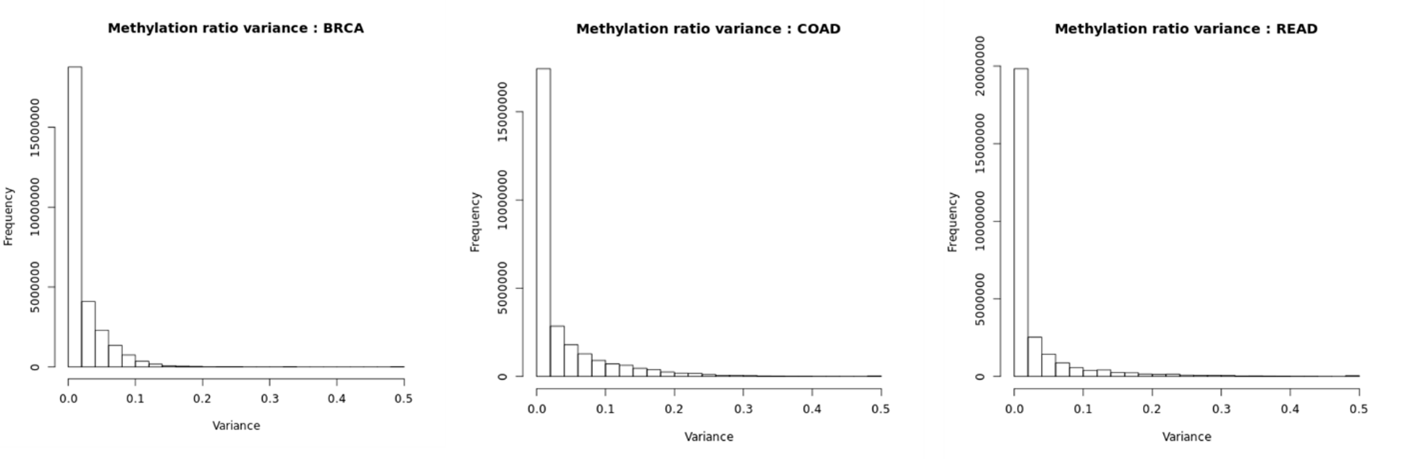

### Supplemental Figure 3

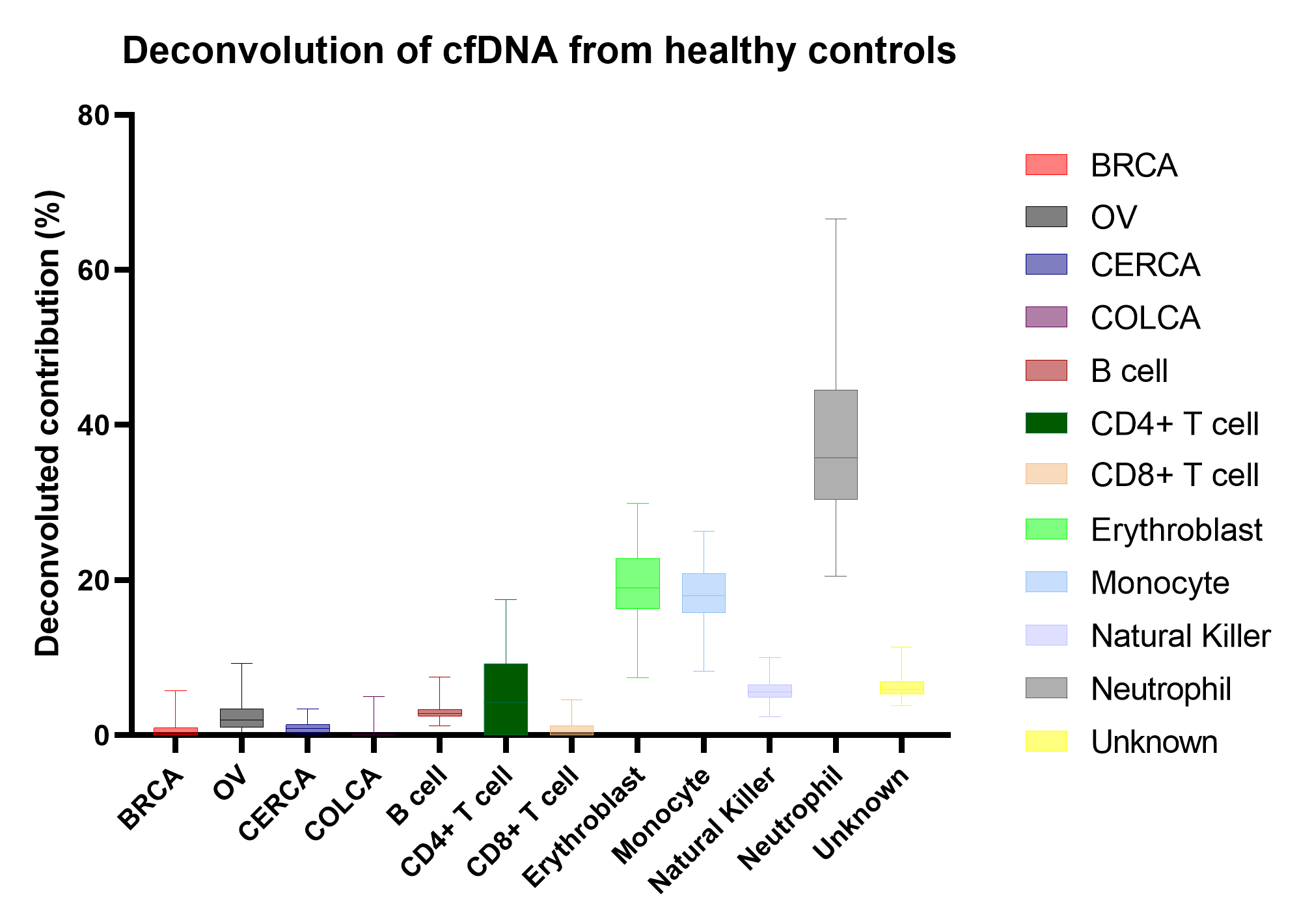
